# Cost-effectiveness of testing for *Mycoplasma genitalium* among men who have sex with men in Australia

**DOI:** 10.1101/2022.08.24.22279191

**Authors:** Jason J. Ong, Aaron G. Lim, Catriona S. Bradshaw, David Taylor-Robinson, Magnus Unemo, Patrick J. Horner, Peter Vickerman, Lei Zhang

## Abstract

**Objectives:** *Mycoplasma genitalium* (MG) disproportionately affects men who have sex with men (MSM). We determined the cost-effectiveness of testing strategies for MG using a healthcare provider perspective.

**Methods:** We used inputs from a dynamic transmission model of MG among MSM living in Australia in a decision tree model to evaluate the impact of four testing scenarios on MG incidence: 1) no one tested; 2) symptomatic MSM; 3) symptomatic and high-risk asymptomatic MSM; 4) all MSM. We calculated the incremental cost-effectiveness ratios (ICERs) using a willingness to pay threshold of $30,000 AUD per QALY gained. We explored the impact of adding an AMR tax (i.e. additional cost per antibiotic consumed) to identify the threshold whereby any testing for MG is no longer cost-effective.

**Results:** Testing only symptomatic MSM is the most cost-effective (ICER $3,677 per QALY gained) approach. Offering testing to all men is dominated (i.e. not recommended because of higher costs and lower QALYs gained compared to other strategies). When the AMR tax was above $150, any testing for MG was no longer cost-effective.

**Conclusion:** Testing only symptomatic MSM is the most cost-effective option even when the potential costs associated with AMR are accounted for (up to $150 additional cost per antibiotic consumed). For pathogens like MG where there are anticipated future costs related to AMR, we recommend models to test the impact of incorporating these costs as they can change the conclusions of cost-effectiveness studies.

**KEY MESSAGES:**

- **What is already known on this topic** - *Mycoplasma genitalium* (MG) is a sexually transmitted pathogen with rising antimicrobial resistance.
- **What this study adds** - This economic evaluation found that testing only symptomatic men who have sex with men (MSM) is the most cost-effective option. When the costs per antibiotic consumed is greater than $150, any testing for MG is no longer cost-effective.
- **How this study might affect research, practice or policy** - Among testing strategies for MSM, testing for MG should be restricted to symptomatic men only.

## INTRODUCTION

*Mycoplasma genitalium* (MG) is a sexually transmitted bacterium that can cause urethritis, cervicitis, endometritis, and pelvic inflammatory disease.^1^ A recent systematic review estimates that MG prevalence is 1.3% (95% CI: 1.0-1.8) among the asymptomatic general population in countries with higher Human Development Index (HDI) levels and 3.9% (95% CI: 2.2-6.7) in countries with lower HDI levels.^2^ Of concern, there is a high and rising prevalence of antimicrobial resistance (AMR) in MG globally.^3^

Studies in men who have sex with men (MSM) have reported a higher prevalence of MG compared to the general population. For example, in a study of 1001 asymptomatic MSM attending a sexual health clinic in Australia, a prevalence of 9.5% was reported.^4^ However, although MG may cause urethritis and proctitis in MSM,^1,5^ there are no reproductive sequelae as in women.^6^ Consequently, besides treating MG in MSM to alleviate symptoms, there is uncertainty over the benefits of testing for MG in achieving population-level control among MSM.

Testing asymptomatic individuals for MG is not recommended in the Australian, European, British and US MG management guidelines.^7-10^ This recommendation is currently based on expert opinion. There is a lack of evidence regarding the benefits of testing and treating for MG among asymptomatic individuals, and no cost-effectiveness analysis has been published.

In this study, we aim to determine the cost-effectiveness of testing for MG using a healthcare provider perspective among MSM living in Australia. We evaluated the cost-effectiveness of testing: 1) no one; 2) only symptomatic MSM (current recommendation); 3) symptomatic and high-risk asymptomatic MSM; 4) all MSM. To account for the potential economic impact of future antimicrobial resistant MG, we also imposed an AMR tax to explore whether this would alter the cost-effectiveness of testing strategies. As the future cost of AMR is complex and uncertain, it is challenging to monetize the direct and indirect consequences of AMR, let alone the costs for developing new antibiotics.^11^ So, we adopted the approach used by the World Health Organization in the 2021 Guidelines for the management of symptomatic sexually transmitted infections, in their development of the vaginal discharge management algorithm.^12^ The AMR tax is applied at the point of antibiotic use, and is a way to incorporate the detrimental effects of AMR into the net present value of the interventions under consideration.^13^

## METHODS

### Costs

A micro-costing approach was used to identify costs from an Australian healthcare provider perspective for testing and managing MG. The direct medical economic costs of testing and management were collated from the Australian Pharmaceutical Benefits Scheme and data from Melbourne Sexual Health Centre (MSHC), the largest publicly funded sexual health centre in Australia. We accounted for costs related to the medical consultation, tests and antibiotics used. The type of antibiotics used to manage MG infection was based on the current management guidelines at MSHC (Table 1) and the European guidelines^10^ (as part of our sensitivity analysis). All costs are reported in Australian dollars (2021 AUD). A gamma distribution was used to characterise the uncertainty for costs (based on +/- 30% of the base-case estimate) in the probabilistic sensitivity analysis.

**Table 1.**
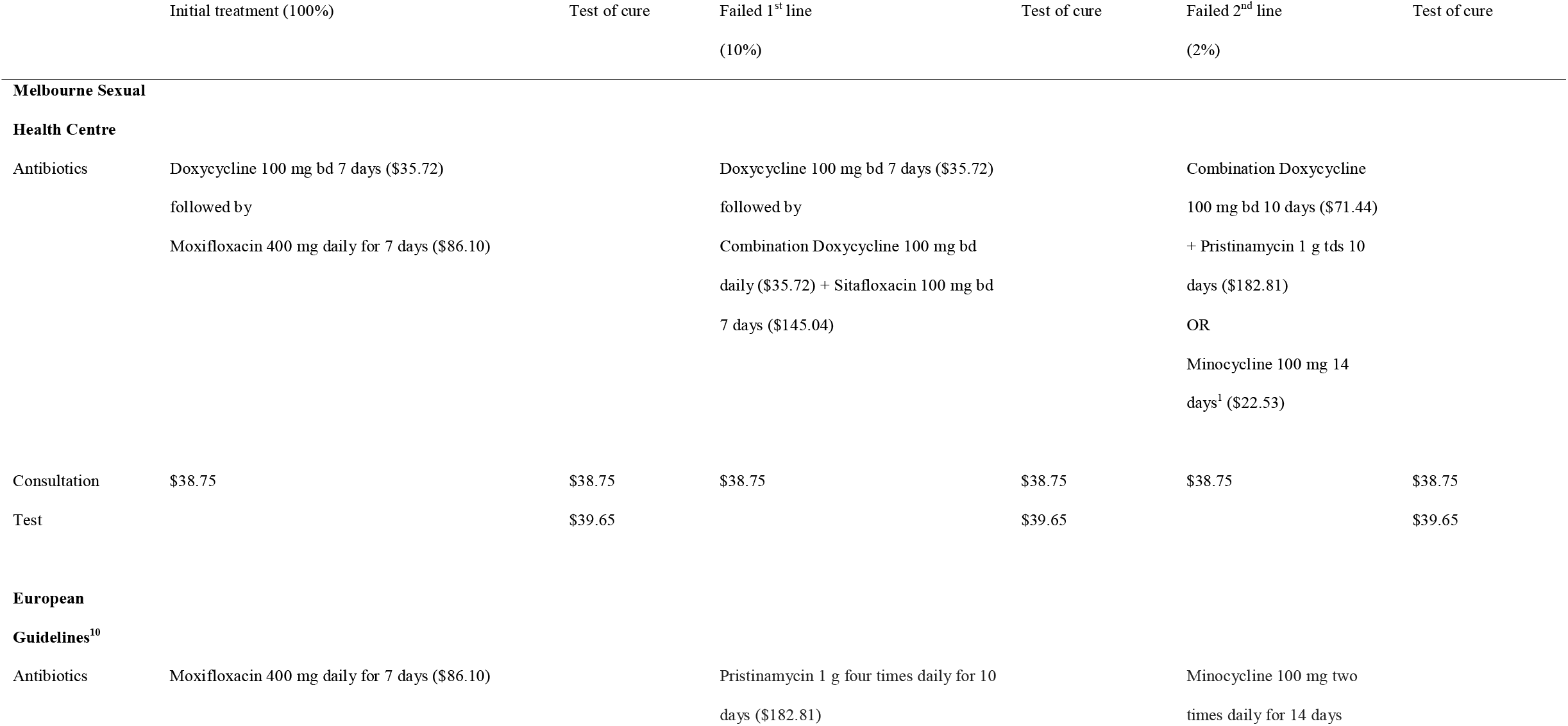
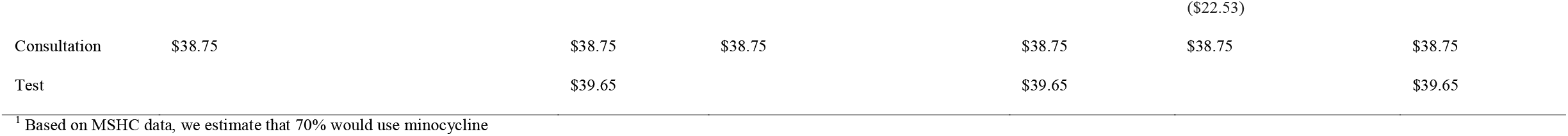
**The direct health costs of managing a patient diagnosed with macrolide-resistant *Mycoplasma genitalium* using antibiotics recommended at Melbourne Sexual Health Centre, and the European guidelines**.

The initial cost of testing includes one consultation and one test. If an individual tests positive for MG, he will incur extra costs related to two additional consultations: one consultation for receiving treatment, and one consultation for the test of cure. The antibiotics prescribed will depend on whether the MG strain is macrolide-susceptible or -resistant (Table 2). We evaluated how adding an AMR tax (ranging from 0 to $200) would affect the cost-effectiveness of testing strategies. There is no single willingness to pay (WTP) threshold in Australia, although there is evidence that medicines are more likely to be recommended for listing with an ICER around $30,000 than above $70,000.^14^ We used the more conservative WTP of $30,000 per QALY gained in the analysis, but also provide the probability of being cost-effective across a range of WTP in the cost-effectiveness acceptability curves.

**Table 2.** Cost (AUD) model input parameters.

| Cost parameter | Cost (\$) | Gamma distribution<br>(alpha, lambda) | Reference and assumptions |
| --- | --- | --- | --- |
| Doctor's consultation | 38.75 | 11.1, 0.29 | PBS<sup>34</sup> |
| Test for MG (including test for macrolide resistance) | 39.65 | 11.1, 0.28 | MBS<sup>35</sup> + \$11 for resistance test<sup>1</sup> |
| Azithromycin 1g stat then 500 mg daily for another 3 days | 67.68 | - | PBS<sup>34</sup> |
| Doxycycline 100 mg twice daily for 1 week | 35.72 | - | PBS<sup>34</sup> |
| Moxifloxacin 400 mg daily for 7 days | 86.10 | - | MSHC<sup>36</sup> |
| Pristinamycin 1g three times a day for 10 days | 182.81 | - | MSHC<sup>36</sup> |
| Minocycline 100 mg twice daily for 14 days | 22.53 | - | PBS<sup>34</sup> |
| Sitafloxacin 100 mg twice daily for 7 days | 145.04 | - | MSHC<sup>36</sup> |
| Cost of antibiotics for treating wild type (MSHC)<sup>2</sup> | 103.40 | 11.1, 0.11 | PBS<sup>34</sup> |
| Cost of antibiotics for treating wild type (European guidelines) | 67.68 | 11.1, 0.16 | <sup>10</sup> |
| Cost of antibiotics for treating resistant MG (MSHC) | 147.45 | 11.1, 0.08 | PBS<sup>34</sup> |
| Cost of antibiotics for treating resistant MG<sup>2</sup> (European guidelines) | 104.83 | 11.1, 0.11 | <sup>10</sup> |

<sup>1</sup> Discussion with local laboratory (September 2021)
<sup>2</sup> Based on Doxycycline 100 mg bd 1 week followed by Azithromycin 1 g stat, then 500 mg daily for another 3 days
AUD = Australian dollars, MBS = Medicare Benefits Schedule; MG = *Mycoplasma genitalium*; MSHC = Melbourne Sexual Health Centre;
PBS = Pharmaceutical Benefits Scheme

### Model

We created a decision tree model in TreeAgePro 2021 (TreeAge Software, Williamstown, MA, USA) for four testing scenarios: 1) no one tested; 2) only symptomatic MSM tested (the current recommendation); 3) symptomatic and high-risk asymptomatic MSM tested; 4) all MSM tested. We defined high-risk men as those who reported more than ten sexual partners in the last six months.^15^ Figure 1 presents the structure of one testing scenario, which is applied to all four scenarios in the final model. We used this decision tree model to calculate the total cost, total QALYs gained and incremental cost-effectiveness ratio for the four testing scenarios using estimates from a dynamic transmission model of MG among MSM living in Australia that we previously developed.^16^ This dynamic transmission model simulated a cohort of MSM who could be infected with wild-type or macrolide-resistant MG. The dynamic model assumes: 1) a closed cohort of MSM (i.e. no transmission between MSM and non-MSM groups, and no births or deaths were incorporated); 2) a treatment effectiveness of 92-95% using resistance-guided therapy;^17^ 3) no transitions between low- and high-risk men, but sexual mixing can occur between the two populations; 4) the transmission rate is independent of symptom status and the type of MG strain; and 5) 77% of those diagnosed would receive treatment.^5^

**Figure 1.**
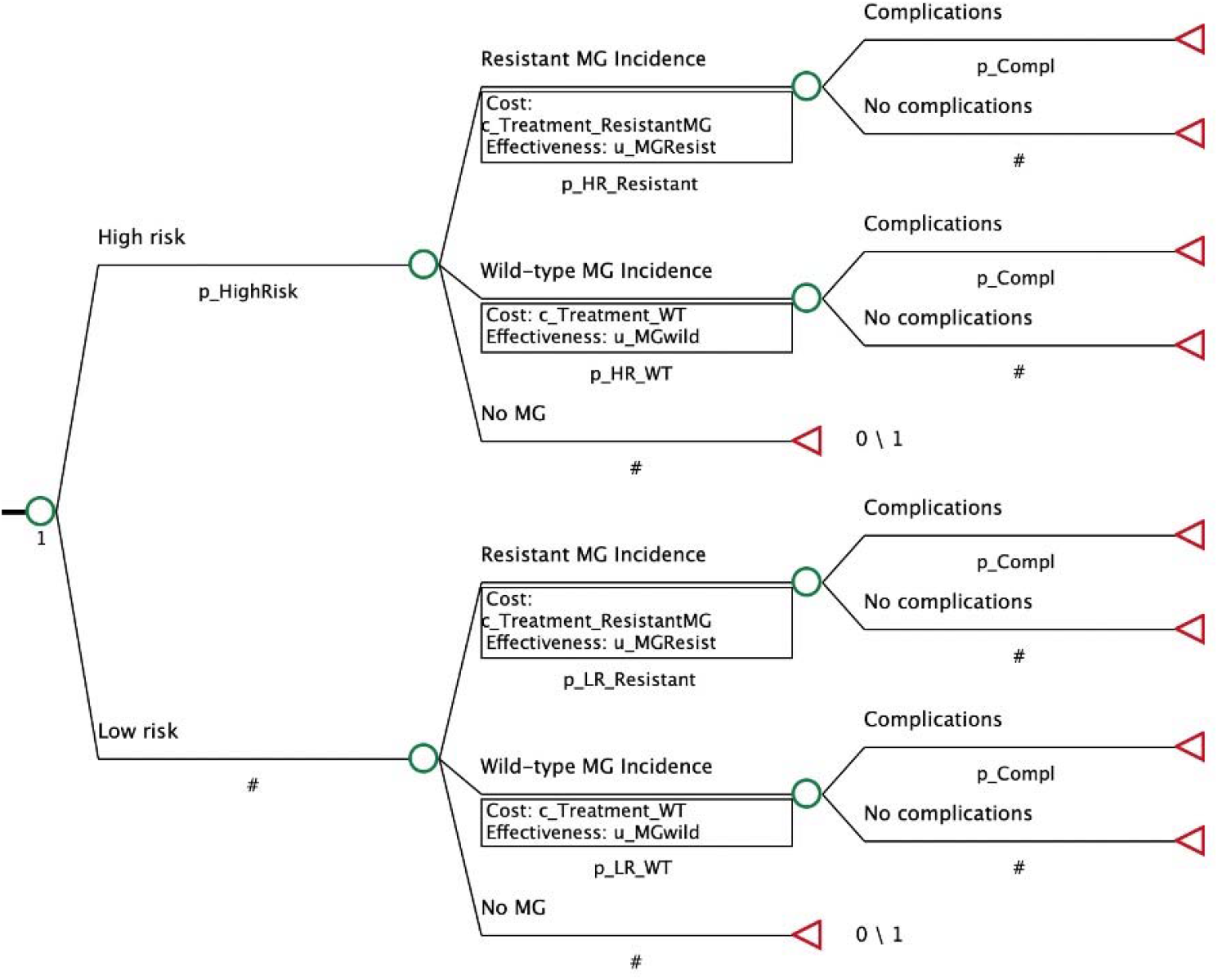
Decision tree model for the cost-effectiveness of testing for *Mycoplasma genitalium* among men who have sex with men. p_HighRisk *=* probability of a men being at high-risk for *Mycoplasma genitalium* (MG) p_HR_Resistant = probability that MG is resistant to macrolides for high-risk MSM p_HR_WT = probability that MG is wild type for high-risk MSM p_LR_Resistant = probability that MG is resistant to macrolides for low-risk MSM p_LR_WT = probability that MG is wild type for low-risk MSM p_Compl = probability of complications related to MG # = 1 – other rates

### Model inputs

There are limited data for utility weights related to MG (eTable 1). For men with complications (defined as ongoing symptoms and anxiety associated with the infection), we assumed a utility weight of 0.96, which was based on 3 months with disutility weight of 0.16 (based on urethritis).^18^ For men receiving a positive MG test with a resistant strain, we assumed 100% had a disutility weight of 0.1 for the first month, 10% with disutility weight of 0.1 in the second month, and 2% with disutility weight of 0.1 in the third month. These proportions were based on the clinical experience of sexual health clinicians at MSHC. For men receiving a positive MG test with a wild-type strain, we assumed 100% had a disutility weight of 0.05 for the first month. The probabilities of men having more than 10 partners in the last 6 months, proportion with ongoing symptoms, and proportion of men screened based on symptoms were derived from unpublished data from MSHC.

### Sensitivity analyses

To examine the impact of parameter uncertainty (eTable 2), we conducted one-way and probabilistic sensitivity analyses and present these as tornado plots and cost-effectiveness acceptability curves, respectively. A Monte Carlo simulation with 100,000 samples was run for the probabilistic sensitivity analysis. In secondary analyses, we tested the effect of a range of AMR tax ($20 to $200) on the cost-effectiveness acceptability curves, and to identify the threshold level of the AMR tax where any testing for MG is no longer cost-effective. We also explored a scenario whereby MG testing is conducted in three anatomical sites (eTable 3).

**Table 3.** **Cost-effectiveness of testing for *Mycoplasma genitalium* among 10,000 men who have sex with men, according to various levels of AMR tax**

|  | Cost | Incremental<br>cost | QALY | Incremental<br>QALY | ICER |
| --- | --- | --- | --- | --- | --- |
| **\$0 AMR tax** | | | | | |
| No one is tested | 0 |  | 9,970 |  |  |
| Testing only symptomatic MSM (current<br>recommendation) | 11,032 | 11,032 | 9,973 | 3 | 3,677 |
| Testing symptomatic and high-risk<br>asymptomatic MSM | 103,495 | 92,463 | 9,975 | 2 | 46,232 |
| Testing all MSM | 1,417,606 | 1,314,111 | 9,961 | -14 | Dominated |
| **\$20 AMR tax** | | | | | |
| No one is tested | 0 |  | 9,970 |  |  |
| Testing only symptomatic MSM (current<br>recommendation) | 15,939 | 15,939 | 9,973 | 3 | 5,313 |
| Testing symptomatic and high-risk<br>asymptomatic MSM | 116,265 | 100,326 | 9,975 | 2 | 50,163 |
| Testing all MSM | 1,463,541 | 1,347,276 | 9,961 | -14 | Dominated |
| **\$50 AMR tax** | | | | | |
| No one is tested | 0 |  | 9,970 |  |  |
| Testing only symptomatic MSM (current<br>recommendation) | 23,300 | 23,300 | 9,973 | 3 | 7,767 |
| Testing symptomatic and high-risk<br>asymptomatic MSM | 135,419 | 112,119 | 9,975 | 2 | 56,060 |
| Testing all MSM | 1,532,443 | 1,397,024 | 9,961 | -14 | Dominated |

AMR = antimicrobial resistance; ICER = incremental cost-effectiveness ratio; MSM = men who have sex with men; QALY = quality- adjusted life years

We report our findings using the Consolidated Health Economic Evaluation Reporting Standards 2022 (CHEERS 2022, Appendix).

## RESULTS

### Primary cost-effectiveness analysis

Table 3 summarizes the total and incremental cost and QALYs gained in a population of 10,000 MSM. It shows that testing all men will always be dominated (i.e. not recommended because of high costs and lower QALYs gained compared to other strategies) regardless of the range of the AMR tax. Testing symptomatic MSM is cost-effective (ICER $3,677 per QALY gained) compared to no testing. eFigure 1 summarizes the results from the probabilistic sensitivity analyses, demonstrating the probability of being cost-effective at the WTP threshold of $30,000 was 40.7% for testing symptomatic MSM only.

### Secondary cost-effectiveness analyses: implementing an AMR tax

eFigures 2 to 6 demonstrates the impact of increasing the AMR tax from $20 to $200. The strategy of not testing anyone becomes most cost-effective when the AMR tax is more than $150.

### Sensitivity analyses

The univariate sensitivity analysis results are shown in eFigures 7 to 9. The ICER for testing only symptomatic men (current recommendation) compared with no testing was most influenced by the probability of resistant MG in low-risk MSM, the AMR tax and disutility of MG complications. However, the ICERs remain below the willingness to pay threshold, suggesting that testing only symptomatic MSM is cost-effective compared to no testing. The ICER for testing only symptomatic men compared with testing both high-risk asymptomatic men and symptomatic men was most influenced by AMR tax, the probability of wild-type MG in high-risk MSM and probability of wild-type MG. The ICER for testing only symptomatic men compared with testing all MSM was most influenced by the probability of resistant MG in low-risk MSM, the disutility of resistant MG and the AMR tax. However, all ICERs remain cost-saving, suggesting that testing only symptomatic MSM will always be cost-saving compared to testing all MSM. Testing three anatomical sites would increase total costs for each scenario but does not change our conclusions (eTable 3).

## DISCUSSION

With increasing access to nucleic acid amplification tests for MG, it is important to ensure recommendations to test for MG are informed by robust evidence, including economic evaluations. Thus, our cost-effectiveness analysis provides evidence for the value in money for various testing strategies for an infection that is predominantly asymptomatic in MSM^19^, but has rising AMR.^3,20^ Our findings strengthen the evidence base for the current international and national guidelines that recommend testing in symptomatic individuals only.^7,8,21^

The current recommendation to test for MG among symptomatic individuals with specific indications is an example of antibiotic stewardship. Overuse of antibiotics has been linked to rising resistance in many pathogens^22^ and specifically for MG, countries with a higher background of macrolide use has been associated with higher prevalence of macrolide-resistant MG.^23,24^ Thus, restricting antibiotics only for symptomatic MSM minimizes antibiotic consumption in a population that already receives far more antibiotics than the general population.^25^ Screening asymptomatic men for MSM is problematic on a number of levels. There are no data to suggest asymptomatic MG in MSM is posing a significant risk to health and, due to rising AMR, management of MG is becoming increasingly complex and costly, with challenges in accessing second- or third-line antibiotics outside of specialist sexual health services. Further, we need to consider the harms associated with screening asymptomatic MSM, including the psychological morbidity of being diagnosed with an asymptomatic infection that is unlikely to cause complications among MSM but is increasingly hard to cure and subjects individuals to repeated and often long courses of antibiotics.

There are increasing reports of the rapid spread of macrolide-resistant MG and the emergence of fluoroquinolone resistance globally.^3^ Where suboptimal treatment regimens are prescribed (particularly in the context of rising macrolide resistance), offering MG testing might lead to increases in macrolide-resistant MG.^16^ This phenomenon is also observed for malaria^26^ and tuberculosis.^27^ Subtherapeutic or subinhibitory antibiotic concentrations can promote the development of AMR in not only the target organism but also in other pathogens and commensals.^28^ Consistent with this, in settings with greater use of macrolides, there is a higher prevalence of macrolide-resistant MG.^29^ This stresses the urgency of implementing resistance-guided therapy for MG to ensure that the most appropriate antibiotic is selected for eliminating the pathogen.^17^

Since the advent of antibiotics, we have observed that the effectiveness of antibiotics generally wanes over time against most pathogens, particularly when there is over-consumption. Thus, economic models should account for costs related to the development and spread of AMR.^30,31^ Currently, very few models explicitly account for the future costs of AMR.^32^ Most studies include direct health costs but do not consider non-health perspectives (e.g. societal impact).^32^ There remains no consensus of how best to capture these costs.

Importantly, future costs should also include treating sequelae that arise from untreatable infections (including prolonged hospitalization), the need to use increasingly toxic and costly antibiotics, and the development of new antibiotics to combat infection. While it may not be feasible to accurately capture these specific cost items, the strength of our approach to use an AMR tax would incorporate these components to explicitly demonstrate the impact of AMR on present decision-making. However, a limitation is that we could not disaggregate the direct and indirect costs associated with AMR, nor account for the unmeasured impact on other bacteria and the microbiome in the setting of antibiotic overconsumption.

The strength of our study is the use of inputs from a dynamic transmission model for MG to evaluate the cost-effectiveness of a variety of testing strategies for MSM. Given the importance of AMR for MG, we also explored the impact of incorporating potential costs of AMR in our models. Our study should be read in light of some limitations. First, our model calculates the incidence at equilibrium with inputs from the transmission dynamic model; thus, we do not have data on how quickly equilibrium is reached. Second, our findings are specifically for MSM and not for heterosexual populations. The disutility from reproductive morbidity associated with MG will need to be accounted for in future studies in heterosexual populations. Third, the AMR profile of MG varies markedly depending on the country and population,^3^ so we recommend context-specific economic evaluations that also account for locally-derived health and AMR costs. Fourth, there is no consensus on an acceptable threshold for an AMR tax, but we recommend that future models incorporate this as part of their sensitivity analyses given AMR’s significant impact on both cost and effectiveness.^33^ Last, due to absence of data, several model inputs were based on assumptions. Our univariate sensitivity analyses highlight the parameters that would most likely change the ICER of the testing strategy, and thus would benefit from more accurate measurements in future models.

In conclusion, our study confirms that the current recommendation for MG testing, i.e. testing only symptomatic MSM, is the most cost-effective option in an Australian context. This conclusion remains robust despite uncertainties in multiple parameters, including accounting for future costs of AMR.

## Data Availability

All data produced in the present work are contained in the manuscript.

## CONTRIBUTORSHIP

JO, LZ conceived the research idea. JO, AGL, PV and LZ constructed the dynamic transmission model and conducted the economic evaluation. JO drafted the first manuscript. All authors contributed to the writing of the paper and approve the final version.

## PATIENT AND PUBLIC INVOLVEMENT

Patients or the public were not involved in the design, or conduct, or reporting, or dissemination plans of our research.

## DATA AVAILABILITY

All data produced in the present work are contained in the manuscript.

