## Supplementary File for "Cost-effectiveness of testing for *Mycoplasma genitalium* among men who have sex with men in Australia"

**eFigure 1 Cost-effectiveness acceptability curve for testing for *Mycoplasma genitalium* among men who have sex with men ($0 AMR Tax)**

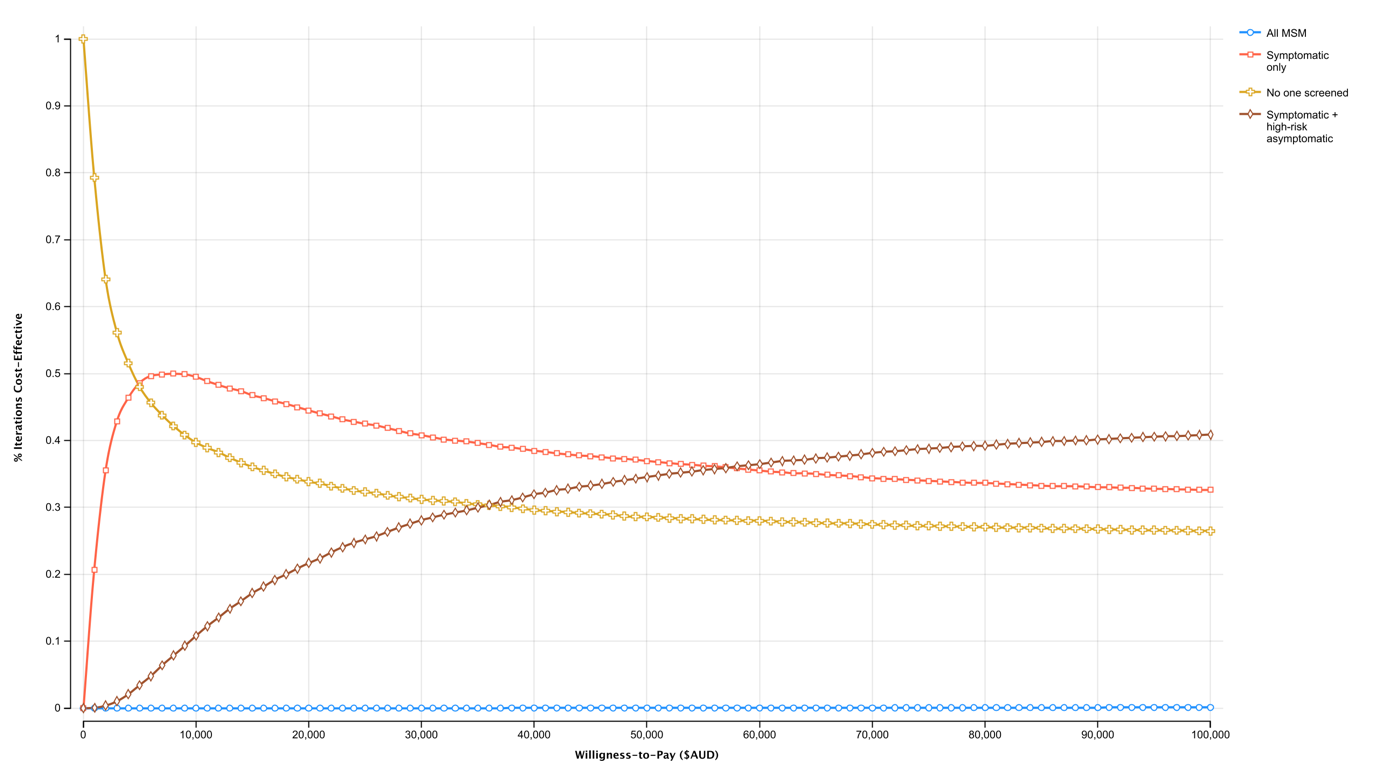

**eFigure 2 Cost-effectiveness acceptability curve for testing for *Mycoplasma genitalium* among men who have sex with men ($20 AMR Tax)**

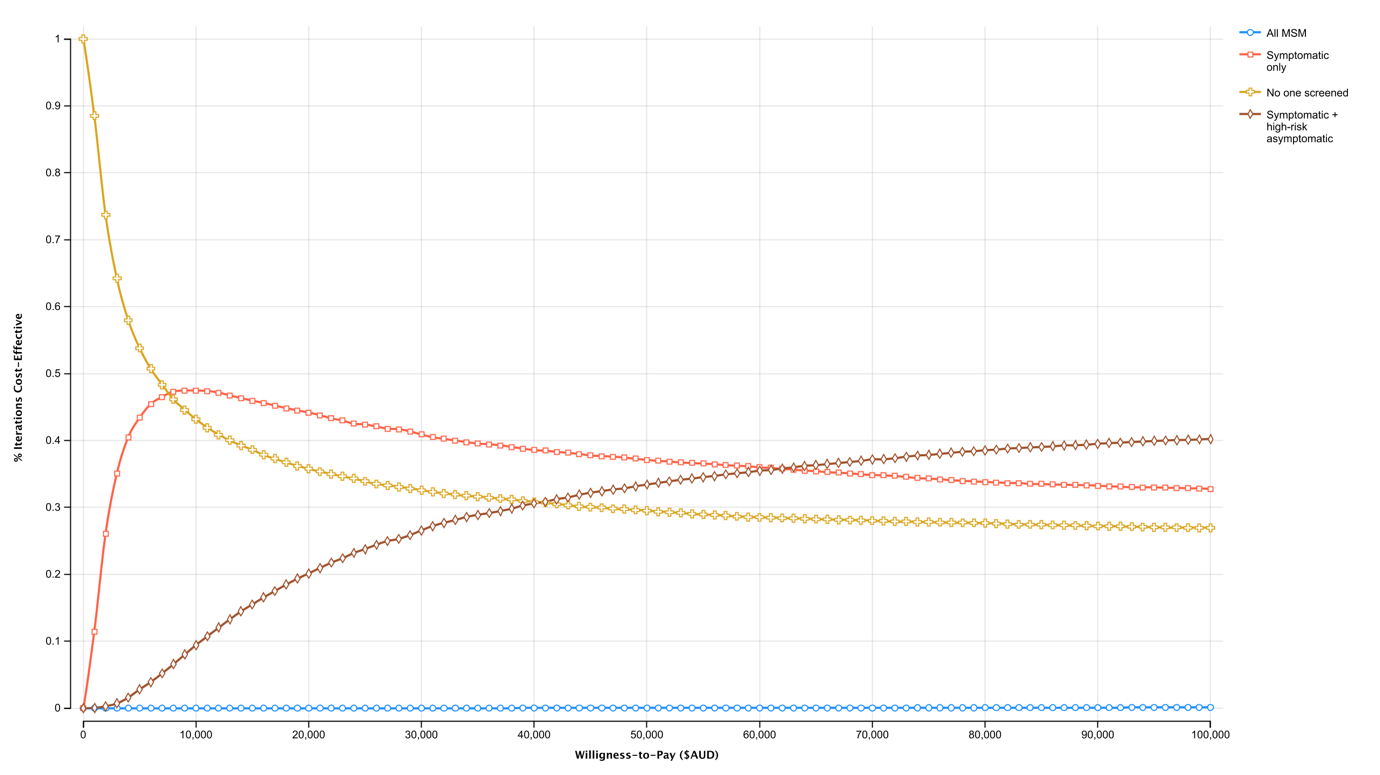

**eFigure 3 Cost-effectiveness acceptability curve for testing for *Mycoplasma genitalium* among men who have sex with men ($50 AMR Tax)**

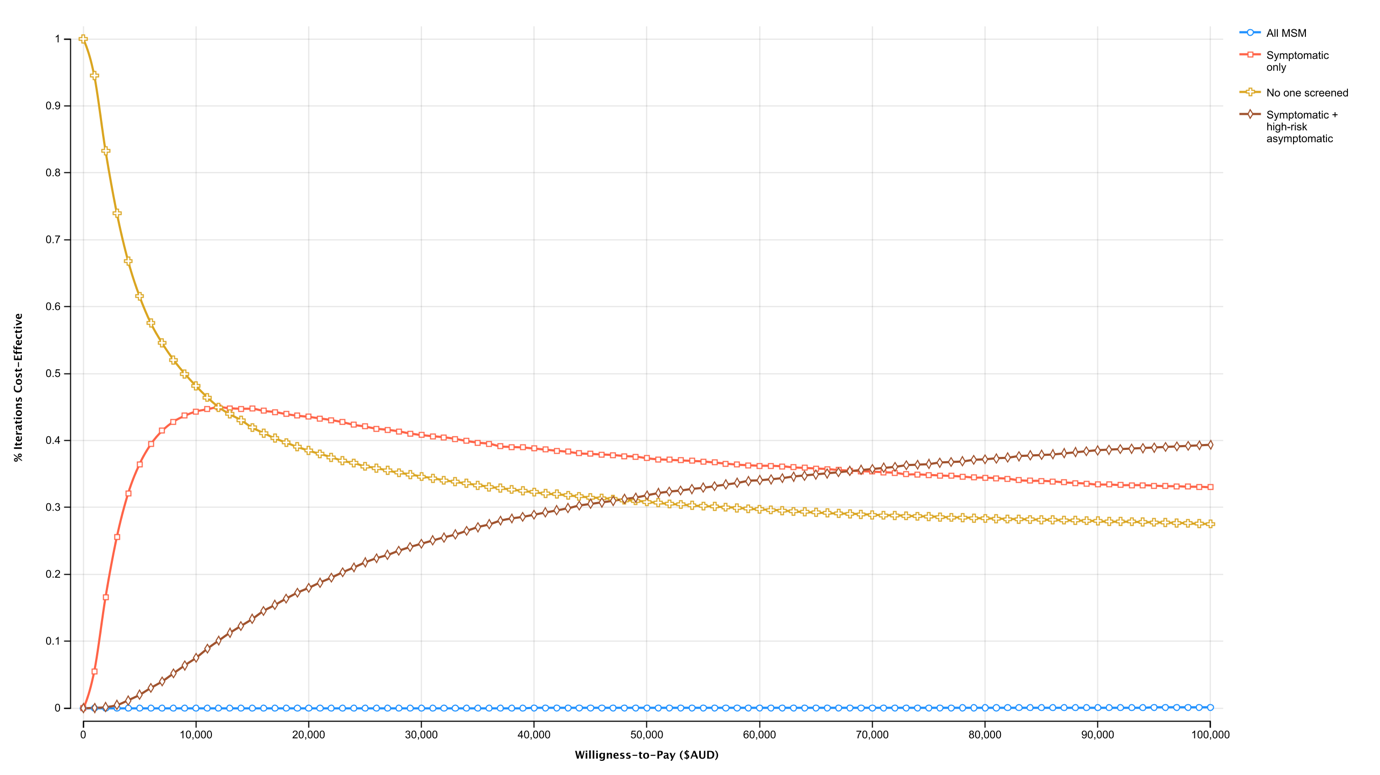

**eFigure 4 Cost-effectiveness acceptability curve for testing for *Mycoplasma genitalium* among men who have sex with men ($100 AMR Tax)**

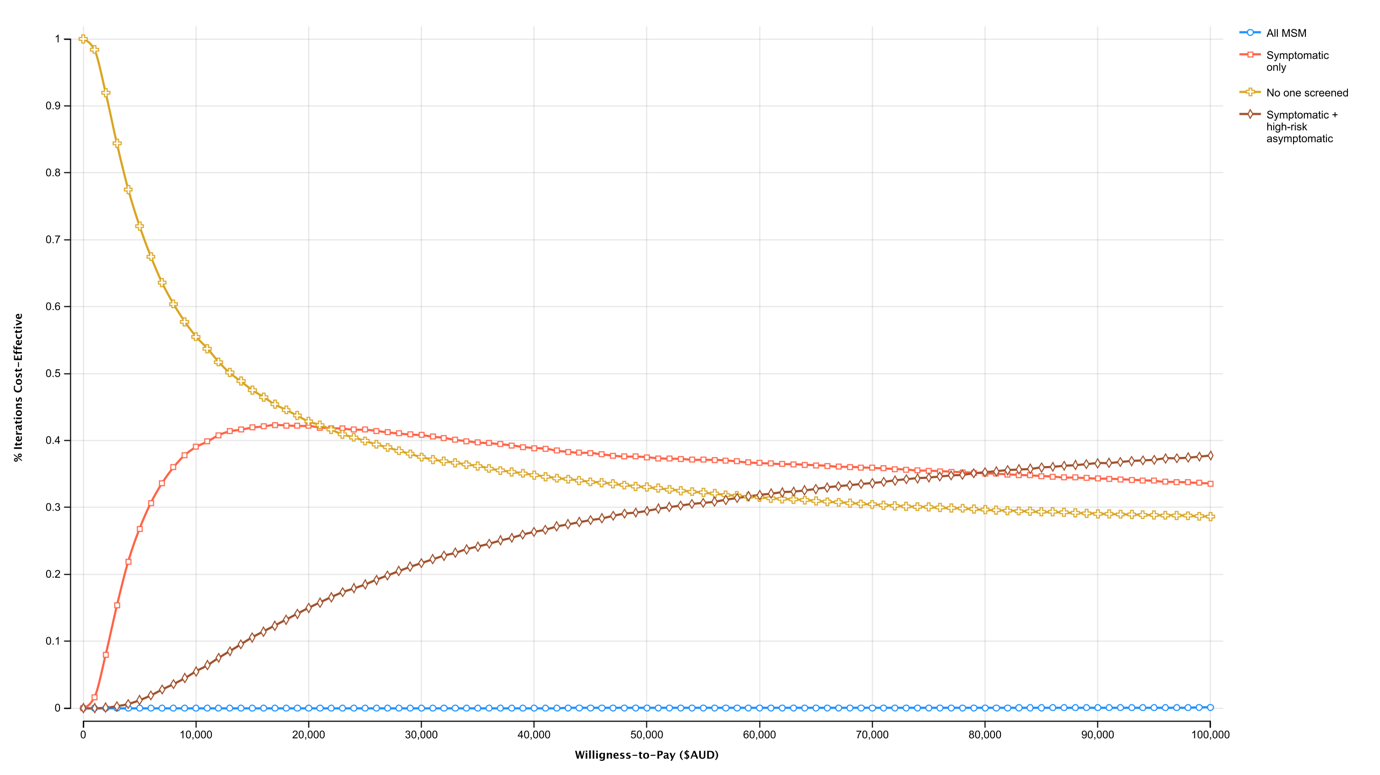

**eFigure 5 Cost-effectiveness acceptability curve for testing for *Mycoplasma genitalium* among men who have sex with men ($150 AMR Tax)**

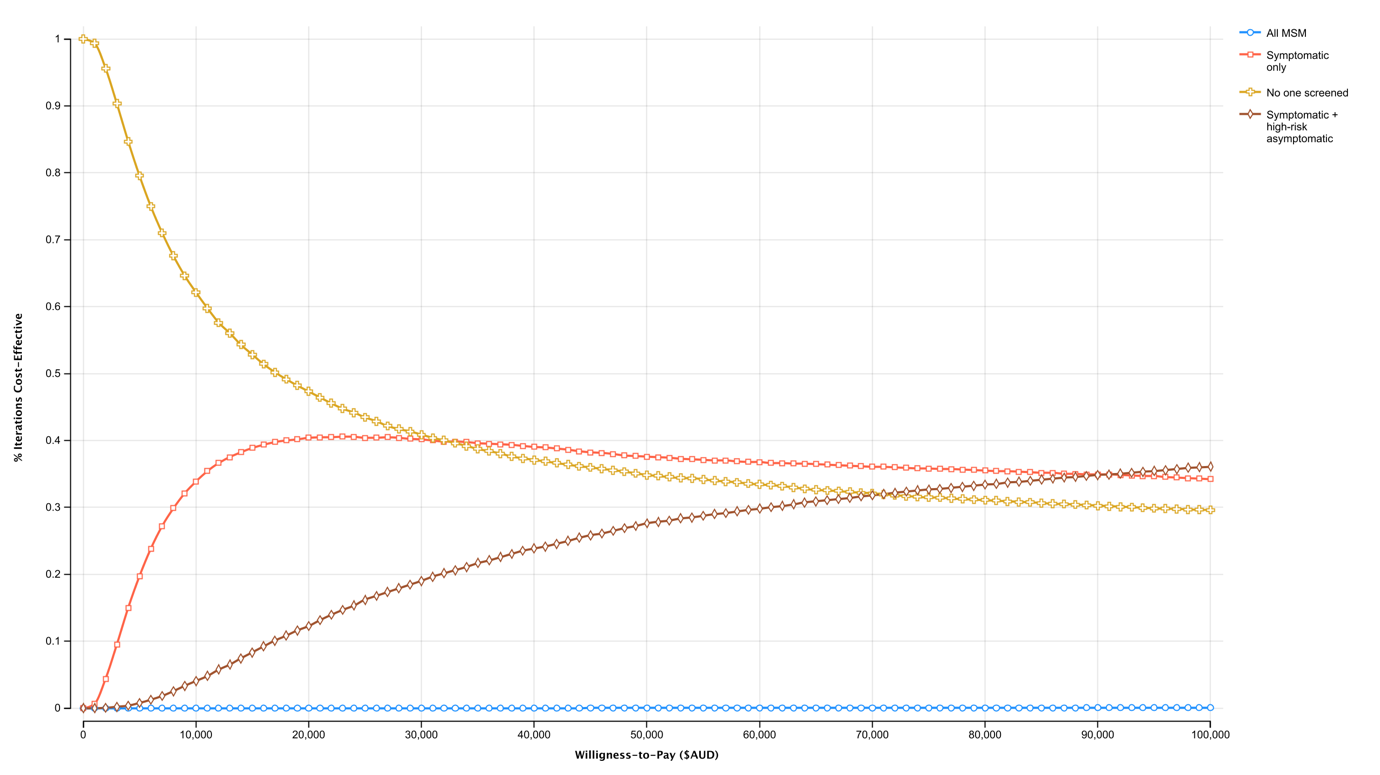

**eFigure 6 Cost-effectiveness acceptability curve for testing for *Mycoplasma genitalium* among men who have sex with men ($200 AMR Tax)**

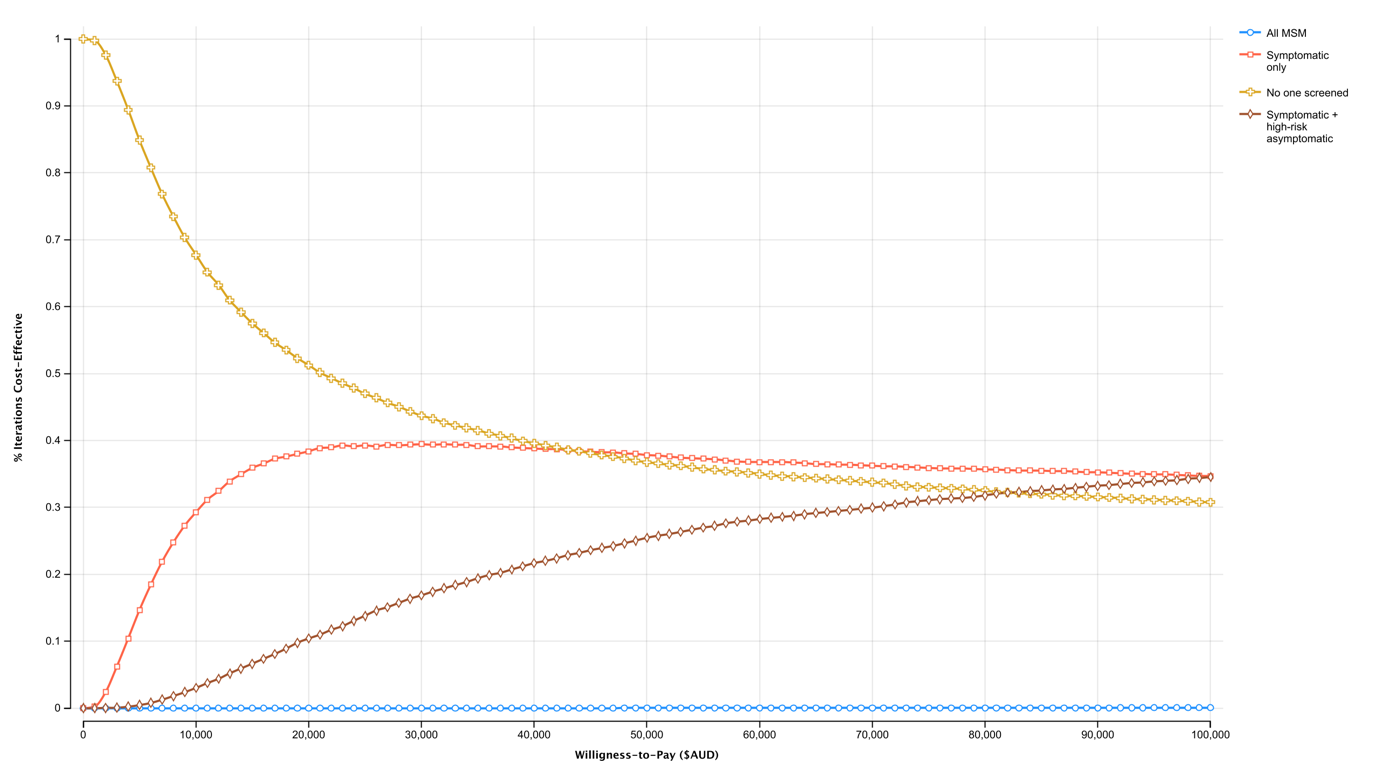

**eFigure 7 Tornado plot of the univariate sensitivity analysis comparing testing symptomatic only vs. no testing**

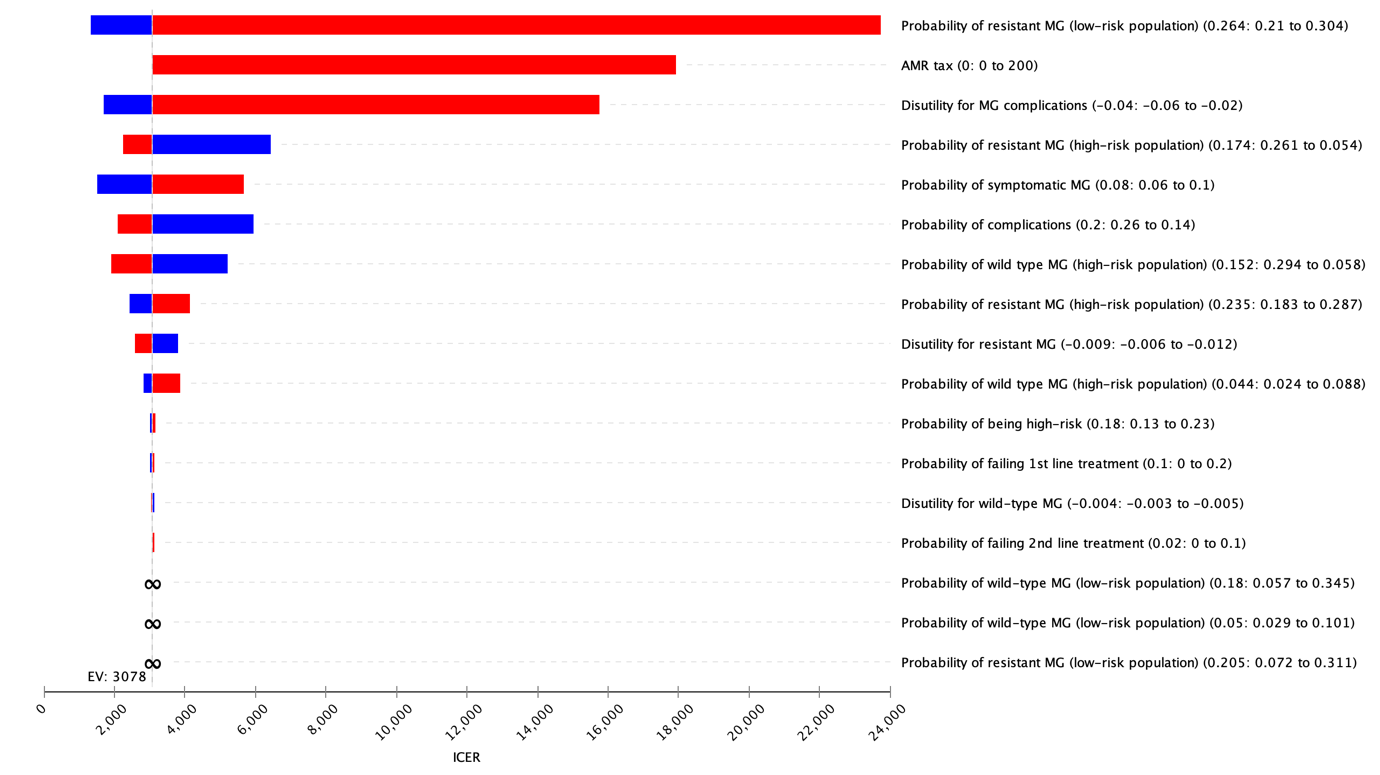

**eFigure 8 Tornado plot of the univariate sensitivity analysis comparing testing symptomatic only vs. testing symptomatic + high-risk MSM**

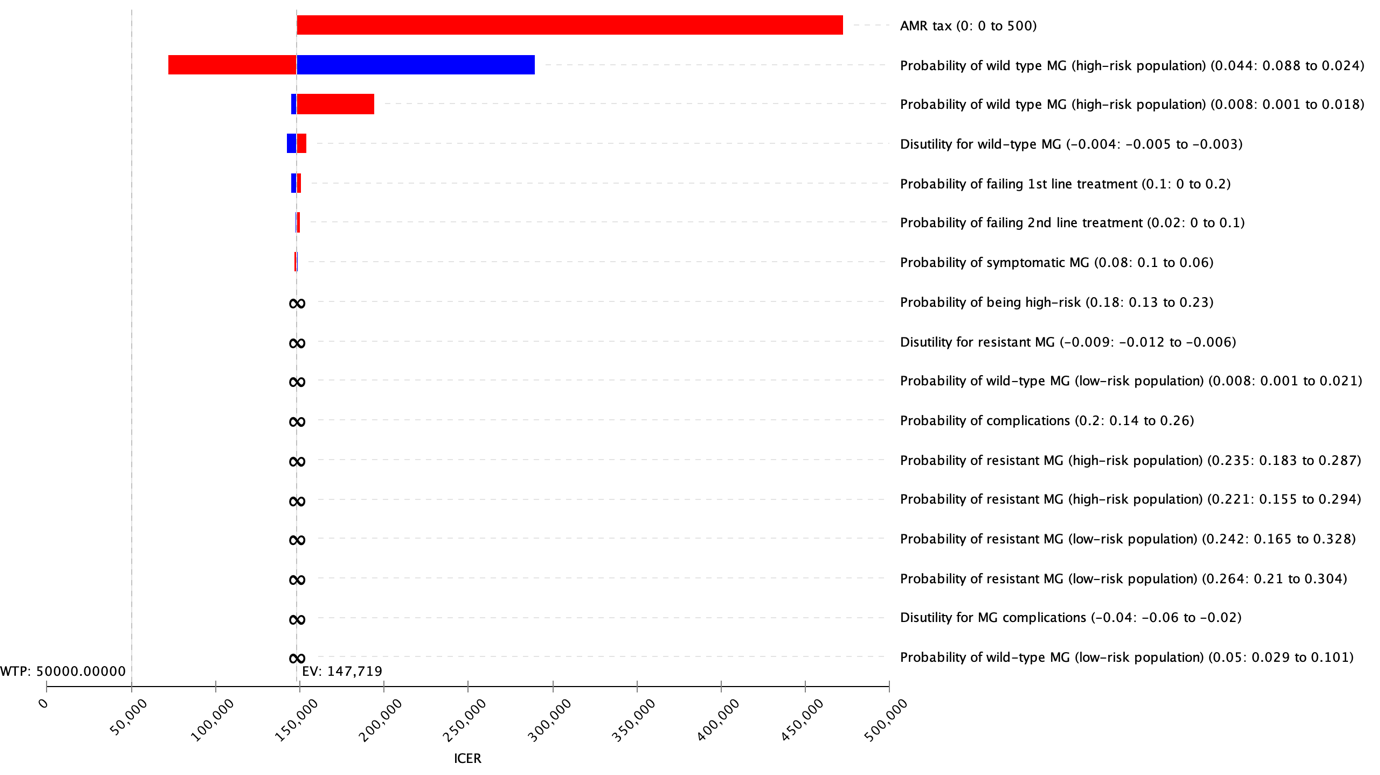

**eFigure 9 Tornado plot of the univariate sensitivity analysis comparing testing symptomatic only vs. testing all MSM**

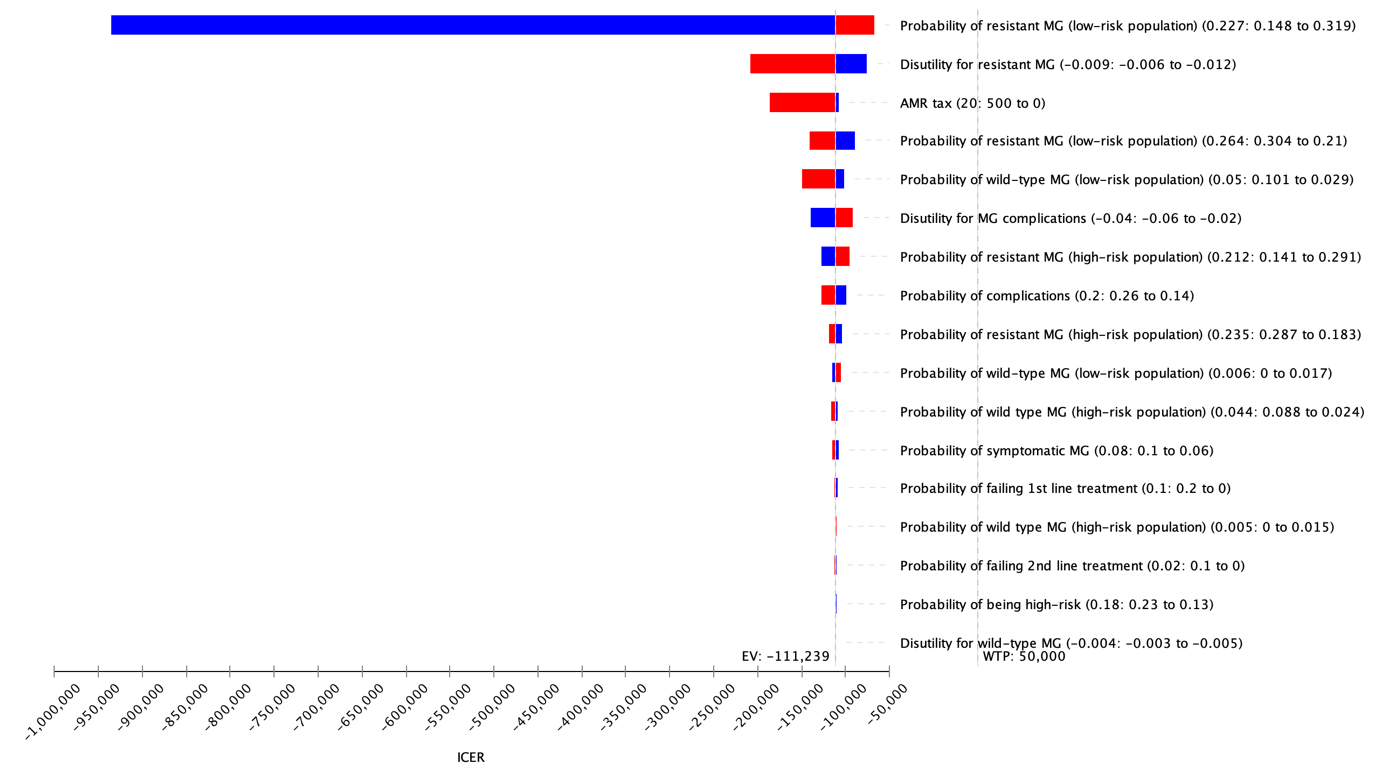

**eTable 1 Utility weights and probabilities for men who have sex with men with *Mycoplasma genitalium***

|  | Utility weight | Beta distribution (alpha, beta) | Reference |
| --- | --- | --- | --- |
| **Health states** |  |  |  |
| Complications | 0.96 | 10.3, 136.4 | 3 months with disutility weight of 0.16 (based on urethritis)^26^ |
| Positive MG test (resistant) | 0.991 | 11.0, 1365.8 | Assuming 100% has disutility weight of 0.1 for 1^st^ month, 10% with additional disutility weight of 0.1 in 2^nd^ month, and 2% with additional disutility weight of 0.1 in 3^rd^ month |
| Positive MG test (wild type) | 0.996 | 11.1, 2754.6 | Assuming 1 month with disutility weight of 0.05 |
| **Probabilities** |  |  |  |
| High-risk MSM | 0.18 | 8.9, 40.7 | Data from MSHC^12^ |
| Complications^1^ | 0.20 | 8.7, 34.8 | Data from MSHC^12^ |
| Symptomatic^2^ | 0.08 | 0.5, 5.9 | Data from MSHC^12^ |

MG = *Mycoplasma genitalium*; MSHC = Melbourne Sexual Health Centre

^1^ This parameter is used in the model to capture the probability of complications related to symptoms and anxiety associated with the infection.

^2^ This parameter is used in the model to determine what proportion of men would be screened based on symptoms.

**eTable 2 Parameters varied for the deterministic sensitivity analyses**

|  | Base-case estimate | Range |
| --- | --- | --- |
| **Cost** |  |  |
| AMR tax | 0 | 0-200 |
| **Probabilities** |  |  |
| *No one is tested* |  |  |
| Wild type, High-risk | 0.152 | 0.058-0.294 |
| Wild type, Low-risk | 0.180 | 0.057-0.345 |
| Resistant, High-risk | 0.174 | 0.054-0.261 |
| Resistant, Low-risk | 0.205 | 0.072-0.311 |
| *Only symptomatic MSM tested* |  |  |
| Wild type, High-risk | 0.044 | 0.024-0.088 |
| Wild type, Low-risk | 0.050 | 0.029-0.101 |
| Resistant, High-risk | 0.235 | 0.183-0.287 |
| Resistant, Low-risk | 0.264 | 0.210-0.304 |
| *Symptomatic and high-risk asymptomatic tested* |  |  |
| Wild type, High-risk | 0.008 | 0.001-0.018 |
| Wild type, Low-risk | 0.008 | 0.001-0.021 |
| Resistant, High-risk | 0.221 | 0.155-0.294 |
| Resistant, Low-risk | 0.242 | 0.165-0.328 |
| *All men tested* |  |  |
| Wild type, High-risk | 0.005 | 0.000-0.015 |
| Wild type, Low-risk | 0.006 | 0.000-0.017 |
| Resistant, High-risk | 0.212 | 0.141-0.291 |
| Resistant, Low-risk | 0.227 | 0.148-0.319 |
| Proportion of high-risk men^1^ | 0.18 | 0.13-0.23 |
| Proportion of men with complications^2^ | 0.20 | 0.14-0.26 |
| Proportion of men who are symptomatic^3^ | 0.08 | 0.06-0.10 |
| **Utility weight** |  |  |
| Complications^1^ | 0.960 | 0.940-0.980 |
| Resistant MG | 0.991 | 0.988-0.994 |
| Wild-type MG | 0.996 | 0.995-0.997 |

^1^ This parameter is used in the model to determine the proportion of men offered testing in two testing scenarios: 1) testing symptomatic MSM only; and 2) testing symptomatic and high-risk MSM

^2^ This parameter is used in the model to capture the probability of complications related to symptoms and anxiety associated with the infection.

^3^ This parameter is used in the model to determine what proportion of men would be screened based on symptoms.

| **eTable 3 Cost-effectiveness of testing for *Mycoplasma genitalium* among 10,000 men who have sex with men, comparing scenario where MG was tested in three anatomical sitesCost-effectiveness of testing for *Mycoplasma genitalium* among 10,000 men who have sex with men, according to various levels of AMR tax Cost-effectiveness of testing for *Mycoplasma genitalium* among 10,000 men who have sex with men, according to various levels of AMR tax**  **Base-case** | Cost | Incremental cost | QALY | Incremental QALY | ICER |
| --- | --- | --- | --- | --- | --- |
| No one is tested | 0 |  | 9,970 |  |  |
| Testing only symptomatic MSM (current recommendation) | 11,032 | 11,032 | 9,973 | 3 | 3,677 |
| Testing symptomatic and high-risk asymptomatic MSM | 103,495 | 92,463 | 9,975 | 2 | 46,232 |
| Testing all MSM | 1,417,606 | 1,314,111 | 9,961 | -14 | Dominated |
| **Testing 3 anatomical sites** | | | | | |
| No one is tested | 0 |  | 9,970 |  |  |
| Testing only symptomatic MSM (current recommendation) | 18,406 | 18,406 | 9,973 | 3 | 6,135 |
| Testing symptomatic and high-risk asymptomatic MSM | 175,276 | 156,870 | 9,975 | 2 | 78,435 |
| Testing all MSM | 2,417,078 | 2,241,802 | 9,961 | -14 | Dominated |
